# Geo-epidemiology of Malaria in Burkina Faso, 2013-2018: a recent re-increase

**DOI:** 10.1101/2021.10.27.21265260

**Authors:** Cédric S. Bationo, Virgil Lokossou, Jordi Landier, Bry Sylla, Gauthier Tougri, Boukary Ouedraogo, Mady Cissoko, Nicolas Moiroux, Jean Gaudart

## Abstract

**Background:** After a global decline, malaria cases re-increases have been shown recently.

The aim of this analysis was to update the epidemiological facies of malaria in Burkina Faso (around 4% of malaria cases worldwide) by estimating weekly malaria incidences at health district levels, from 2013 to 2018, associated to environmental and meteorological factors.

**Methods:** Malaria cases and deaths weekly reports were extracted from the National Malaria Control Program for each health district from 2013 to 2018. Population data were extracted from the reports of the national statistics council. Environmental data were collected through remote sensing.

After estimating incidence through time and space, trend was assessed by an additive decomposition of the incidence and malaria seasons of transmission was estimated by change point analysis (PELT algorithm).

Incidence maps for each year of the study period were assessed to highlight spatial variability through years. Maps of rainfall, temperature, and vegetation (NDVI) were produced to characterize the health district environmental variability.

**Results:** In 2013, 775 cases /100,000 inhab.week were observed in average, and remain roughly constant until 2015. Malaria re-increased from 2016, reaching 2428 cases /100,000 inhab.week in 2018.

From 2013 to 2016, two transmission periods were observed: low from January to July (included) and high from August to December (included).

From 2017 to 2018, an intermediate transmission period from mid-November to early January intercalated between the low transmission period from mid-February to early June and the high transmission period from July to late December

From 2013 to 2015, the most affected districts were located in the center and central-eastern part of the country. From 2016 to 2018 all health districts, except those in the Sahel region, were affected with at least 45,000 cases per 100,000 persons/year. This south-to-north gradient was also observed with rainfall, temperature and NDVI.

**Conclusion:** Malaria incidences re-increased through years and across the country since 2016. But no modification of the environmental factor variability was observed during the same period, in time or space. The re-increase of malaria in Burkina Faso could be due to a real increase of the disease, or to a better access to diagnostic and cure, together with the development of the epidemiological information system.

## Introduction

Malaria is a parasitic disease transmitted by the female Anopheles mosquitoes that remains the most deadly vector-borne disease globally ^1^. While causing a sustained burden in endemic countries, malaria case and infection patterns vary greatly across regions, seasons and from one year to the next^2–4^.

The number of malaria cases was estimated at 229 million in 2019 with more than 94% of cases and deaths localized in the World Health Organization (WHO) African Region^5^. Burkina Faso accounts for 4% of malaria cases worldwide, displays a strongly seasonal influence, and associations with weather parameters have been quantified^5^. Variations in weather conditions and their impact influence the incidence of malaria by affecting its timing and intensity. This phenomenon has been observed in studies in Burkina Faso and Sénégal ^6–8^. Growing evidence regarding the local variations of malaria transmission intensity warrants the adaptation of control measures to the local epidemiological context ^9^.

WHO Malaria Control Program 2016-2030 recommends that National Malaria Control Programs (NMCPs) to tailor their malaria control based on analysis of past and contemporary data, risk factors, and the environment ^10^. Malaria risk mapping using robust approaches and spatial-temporal analyses that consider environmental and meteorological data are therefore necessary to assess the impact of control and identify areas where malaria control strategies need to be adapted ^11–13^. These techniques will then allow the development of weather-based early warning systems able to predict seasonal variations, and to trigger the timely reactive deployment of preventive measures by health authorities ^7,14^.

The objective of this study was to update the epidemiological facies of malaria in Burkina Faso according to the recent re increase in the incidence. We estimated incidence and mortality rates, and analysed the main environmental indicators influencing the transmission of the disease.

## Materials and methods

### Study area

In 2018, Burkina Faso had a population of 20 244 079 and a population density of 72.2 inhabitants/km ^15^. The country is spread over 3 climatic zones: in the north (Sahelian zone), rainfall is less than 600mm/year. While the centre (northern Sudanese) zone receives 600-900mm/year, rainfall in the southern (southern Sudanese) zone exceeds 900mm/year ^16^.

The health district being the operational entity of the national health system ^17^, it will be the spatial unit in our study (Figure 1). In 2018, in average a health district had a median of around 255000 habitants with an interquartile range equal to 177331

**Figure 1:**
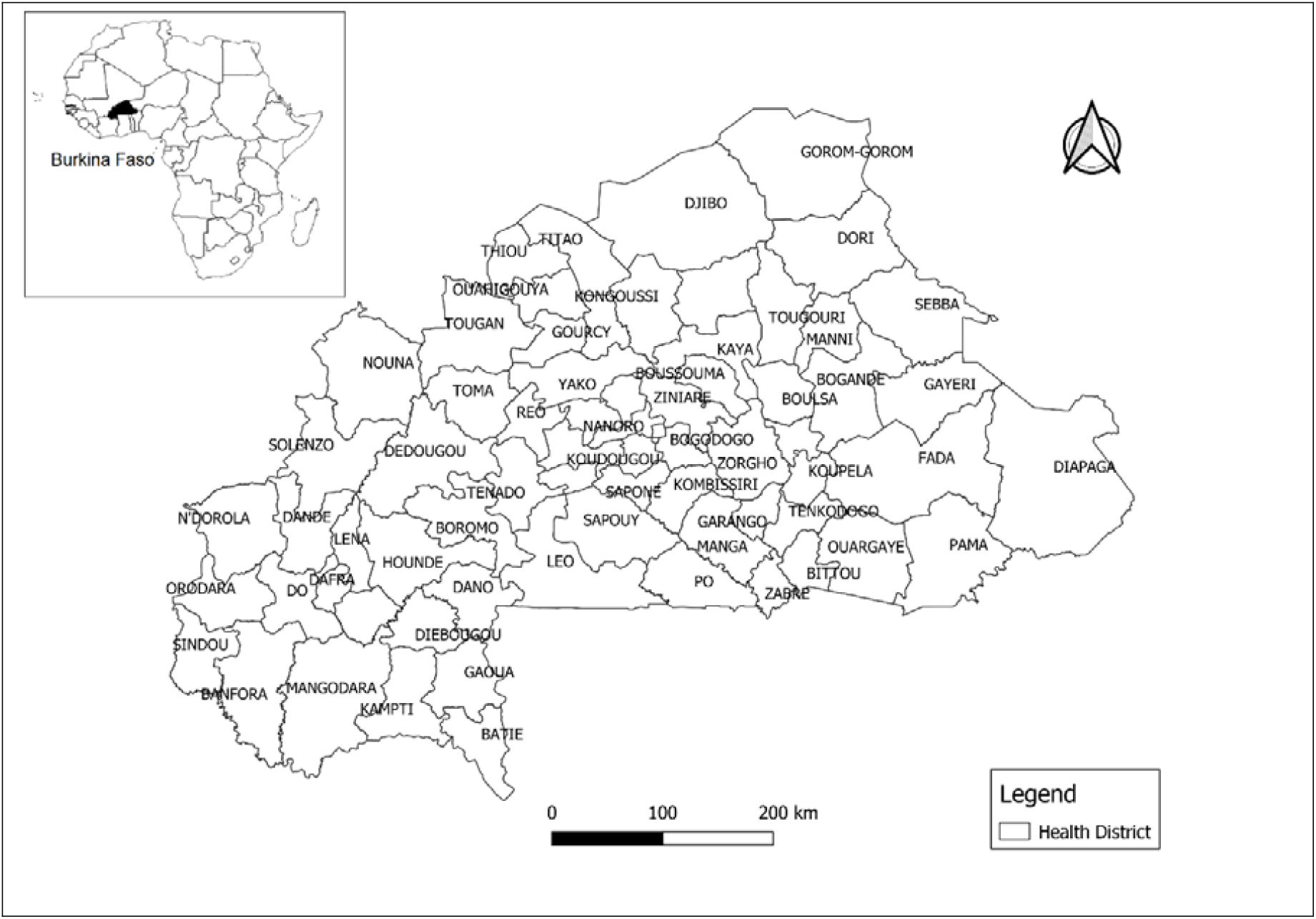
Burkina Faso Map showing the boundaries of the 70 health districts.

### Malaria cases, deaths and population data

A malaria case is defined as a person experiencing fever with a positive rapid diagnostic tests or thick blood smears. Malaria cases and deaths data were obtained from the National Malaria Control Program. Indeed, the national epidemiological surveillance includes an early warning system with priority diseases including malaria. This surveillance is carried out to collect cases and deaths data on a weekly basis, transmitted from the regional to the national level.

We have therefore recovered all malaria cases and deaths in Burkina Faso by week and by health district from January 2013 to December 2018. Yearly estimated population data were extracted from the reports of the national statistics council for the years 2013 to 2018 ^18^.

### Meteorological data

Meteorological data were collected for each health district from remote sensing using satellites through Google earth engine ^19^. The recovered data over the period January 2013-December 2018 are daily precipitation (Climate Hazards Group InfraRed Precipitation with Station Data with, spatial resolution: 0.05 arc degrees), average daily temperature, maximum daily temperature, minimum daily temperature (Latest climate reanalysis produced by ECMWF / Copernicus Climate Change Service, spatial resolution: 0.5 × 0.625°) and the 16 days normalized difference vegetation index (MODIS Terra Vegetation Indices 16-Day Global, spatial resolution: 1000 meters). These data were then aggregated on a weekly basis. We performed linear interpolation to estimate weekly data for the normalized difference vegetation index (NDVI).

### Geographic data

Shapefiles of Burkina Faso Health districts were extracted from the GADM (version 3.6, Davis, CA, USA) Center for Spatial Sciences at the University of California, Davis and Open Street Map ^20^ websites.

### Statistical analysis

#### Temporal analysis

We used the number of weekly malaria cases nationwide and assumed a constant yearly population to estimate the overall incidence time series (2013-2018), estimating trend and seasonality.

Missing case data (weeks 12, 17, and 33 in 2015 and weeks 11, 12, 14, 18, and 38 in 2016) were imputed using linear interpolations. We used an additive decomposition to highlight the stable seasonality. The decomposed curves were smoothed using the LOESS (locally estimated scatterplot smoothing) method. LOESS regression is a non-parametric approach that uses a locally weighted regression to fit a smooth curve across the points of a scatterplot ^21^. We also determined the different transmission periods (low, intermediate and high) through a change point analysis of the mean using the Pruned Exact Linear Time (PELT) algorithm ^22^.

The change point analysis is a method applied on a series of time-ordered data to detect whether changes have occurred, determining the number of changes and estimating dates of changes ^23–25^

#### Data description and mapping

As the impact of meteorological data on malaria incidence being a well-documented phenomenon^7,26,27^, we then superimposed malaria incidence, rainfall and temperature data (mean, minimum and maximum) to assess relationships between incidence variations and meteorological factor, and lags between the time series.

We observed the trend of malaria incidence, incidence density and mortality rates over the years to compare their dynamics.

Finally, we produced choropleth incidence maps at the health district level for the study period in order to highlight the evolution of incidence across the country and possibly observe trends in spatial variations over years. We also produced maps of the averages of rainfall, temperature, and normalized difference vegetation index (NDVI) data over the entire study period to characterize the district environment.

### Software and packages

The statistical analyses were carried out using the software R version 3.6.1 (R Development Core Team, R Foundation for Statistical Computing, Vienna, Austria) ^28^, with the following packages: *imputeTS, ggplottimeseries* ^29^, *tseries* ^30^. The spatial analysis was performed using version 3.10 of the QGIS software (2019, QGIS Development Team) and R.

## Results

### National data description

The national malaria control program recorded 55 417 532 cases of malaria from January 2013 to December 2018 for a population that grew from 17 322 796 in 2013 to 20 244 079 inhabitants in 2018. The median of malaria incidence was 739.06 cases per 100,000 inhabitants/week over the entire period (range 294.46-2427.85). The highest incidences for the years 2013 to 2015 were observed between late July (30th week) and early November (45th week). During the years 2016 to 2018, high incidences were observed between mid-June (25th week) and early November (45th week) (Figure 2).

**Figure 2:**
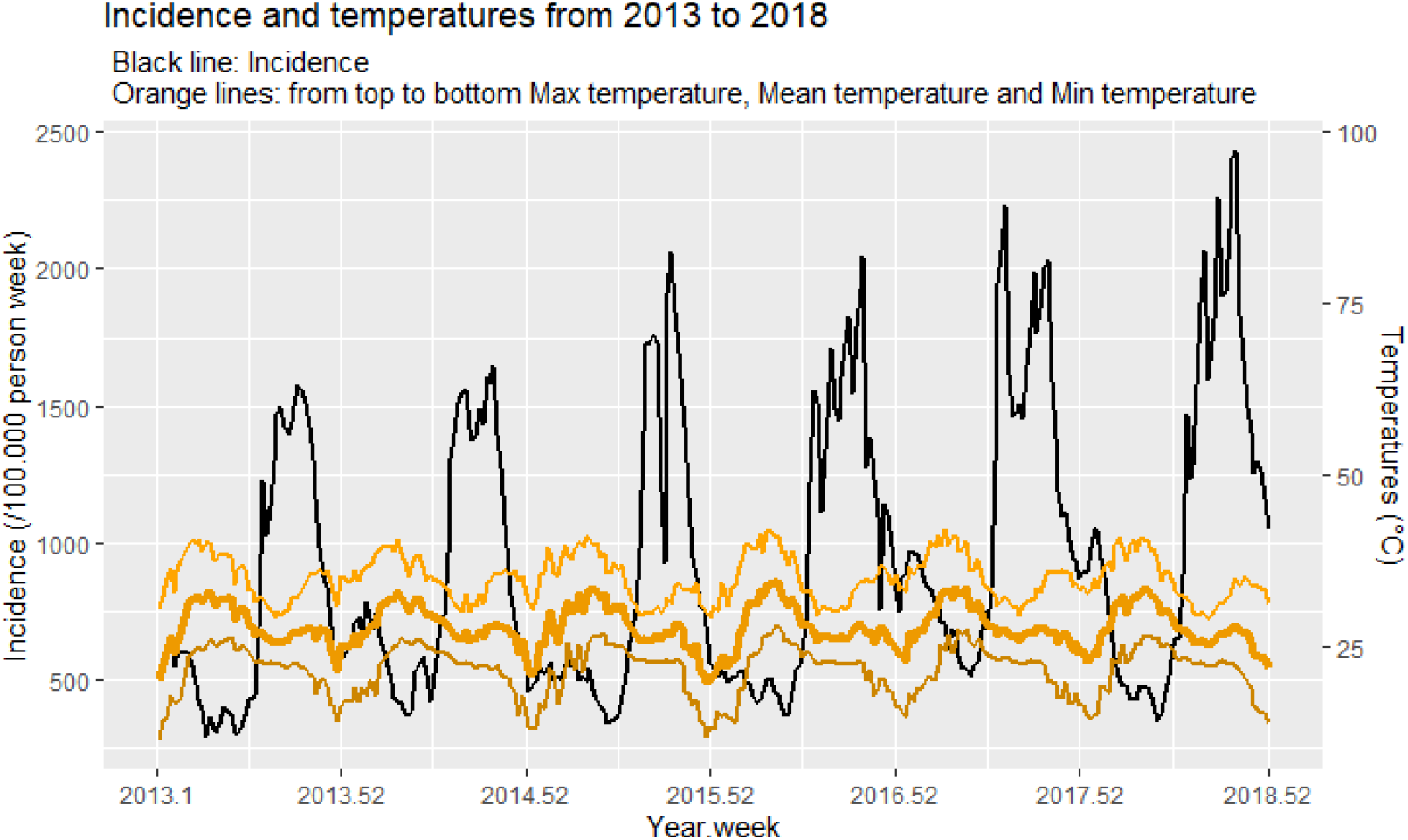
Superposition of malaria incidence and temperature (mean, minimum, and maximum)

Rainfall ranged from 0 to 111.6 mm per week with the highest rainfall recorded between late June and mid-September, and temperatures (mean, minimum and maximum) varied from 11.2 (minimum of minimum temperatures) to 41.9 (maximum of maximum temperatures) with almost constant amplitudes between them (Figure 2 and 3, Table 1). NDVI varied from 0.2 to 0.6 per week (Table 1) with the highest indices recorded between mid-July and mid-October (rainy season).

**Figure 3:**
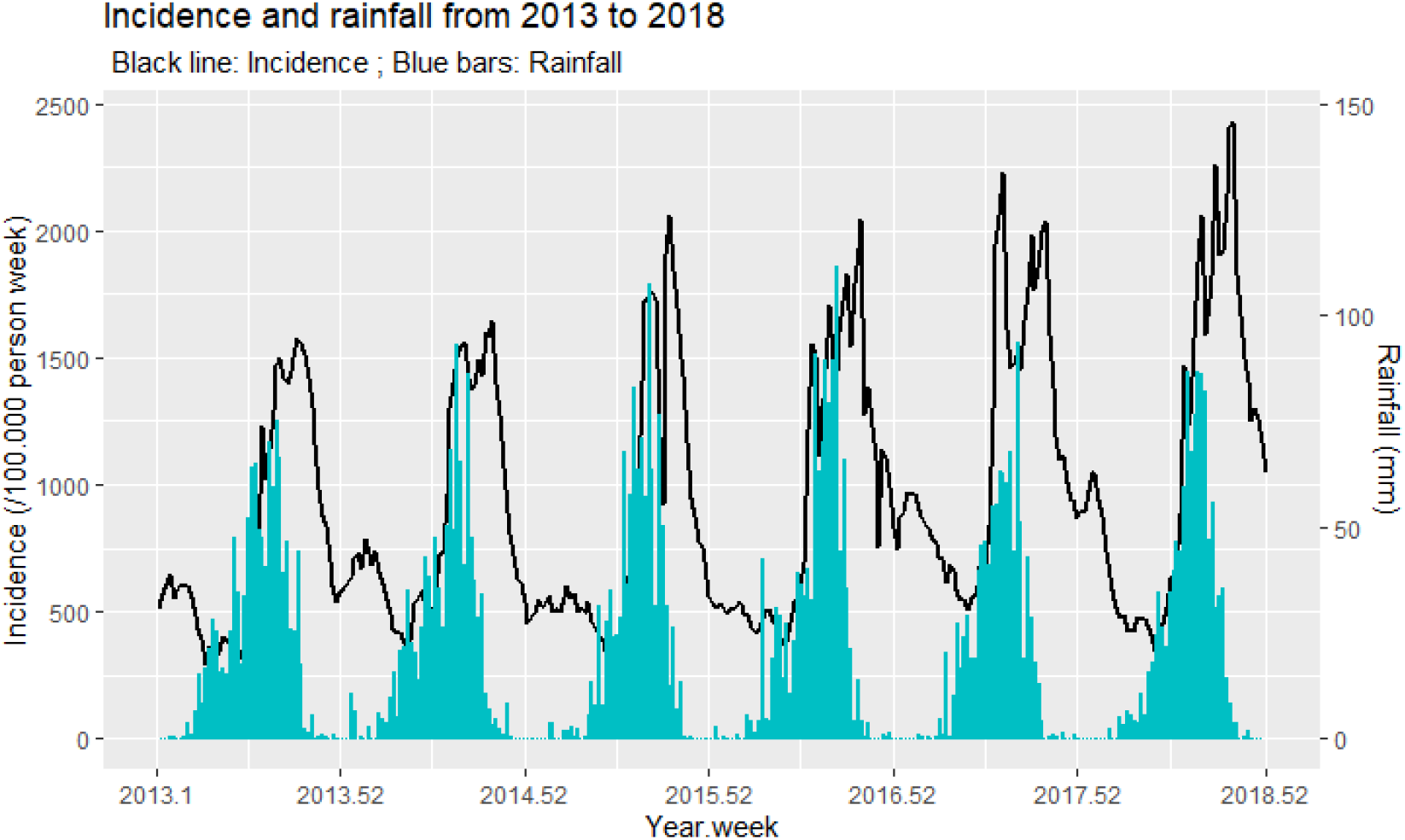
Superposition of malaria incidence and temperature (mean, minimum, and maximum)

In addition to that from 2013 to 2016 we observed two peaks in the incidence density; the first and high one around 500 cases per 100,000 inhabitants/week and the second one around 1500 cases per 100,000 inhabitants/week (Figure 4).

**Figure 4:**
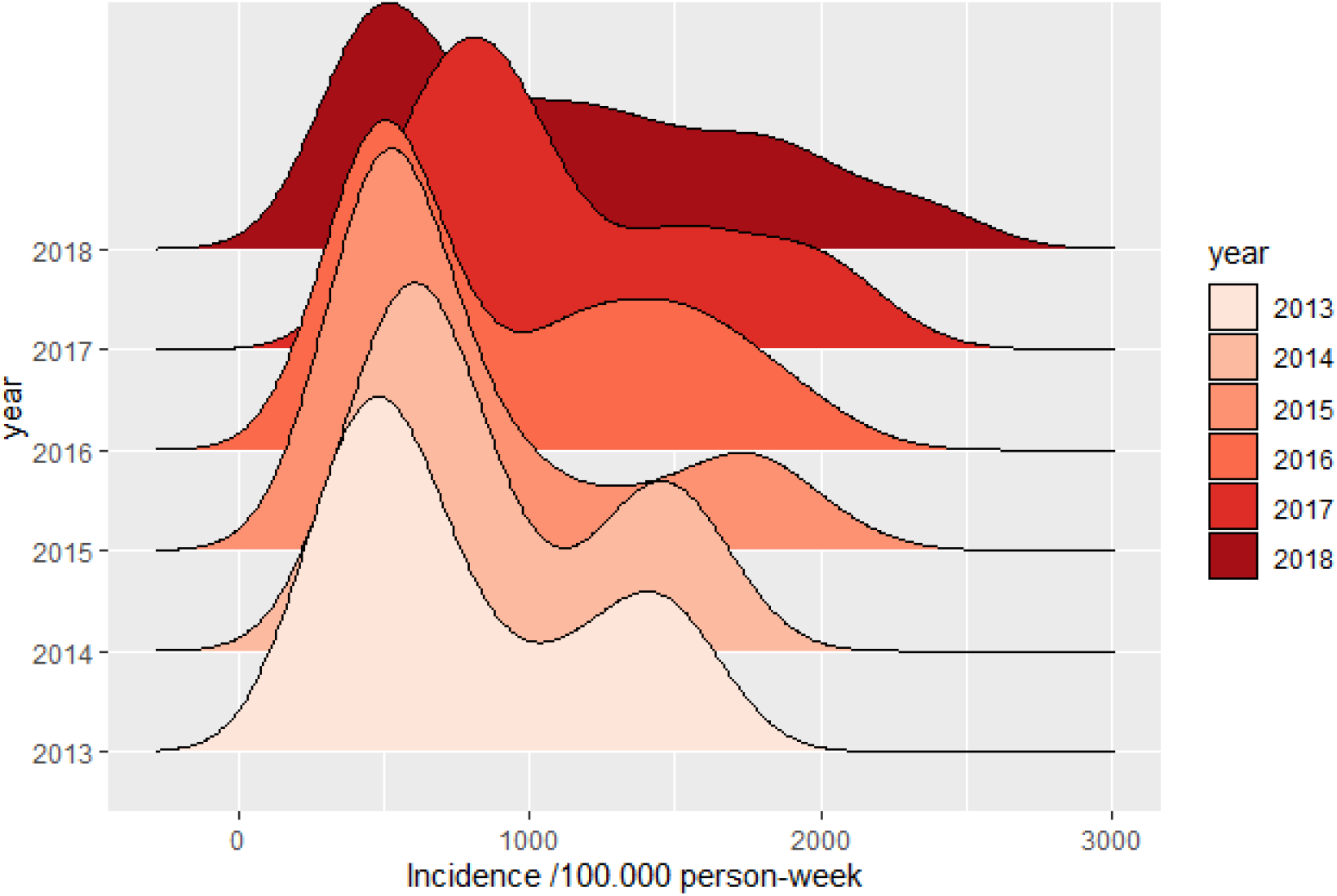
Malaria incidence density from 2013 to 2018

**Table 1:** Lists of weather variables, their abbreviations and weekly descriptive statistics. Var: Variables; St. Dev: Standard deviation; Min: Minimum; Pctl(25): First quartile; Pctl(75): Third quartile; Max: Maximum

| Variable | Mean | St. Dev. | Min | Pctl(25) | Median | Pctl(75) | Max |
| --- | --- | --- | --- | --- | --- | --- | --- |
| Precipitation (mm) | 21.3 | 25.8 | 0 | 0.4 | 8.3 | 35.4 | 111.6 |
| Mean temperature (°C) | 27.7 | 3.0 | 19.8 | 25.9 | 27.3 | 30.2 | 34.3 |
| Maximum temperature (°C) | 34.9 | 3.3 | 29.0 | 32.2 | 34.7 | 37.5 | 41.9 |
| Minimum temperature (°C) | 21.5 | 3.6 | 11.2 | 19.0 | 22.5 | 24.2 | 28.1 |
| Normalized difference vegetation index | 0.3 | 0.1 | 0.2 | 0.2 | 0.3 | 0.4 | 0.6 |

After superimposing malaria incidence and meteorological variables (cumulative weekly rainfall, average of weekly average temperatures, and mean of weekly minimum and maximum temperatures), malaria incidence seemed to increase, but meteorological factors seemed stable over years. (Figure 2 and 3).

### Temporal analysis

The decomposition in trend and season exhibits two clear phases in the time series: a stable phase with a constant incidence (around 800 cases / 100,000 person-years) from 2013 to 2015, followed by an increase phase following a nearly linear trend from 2016 to 2018 (Figure 5).

**Figure 5:**
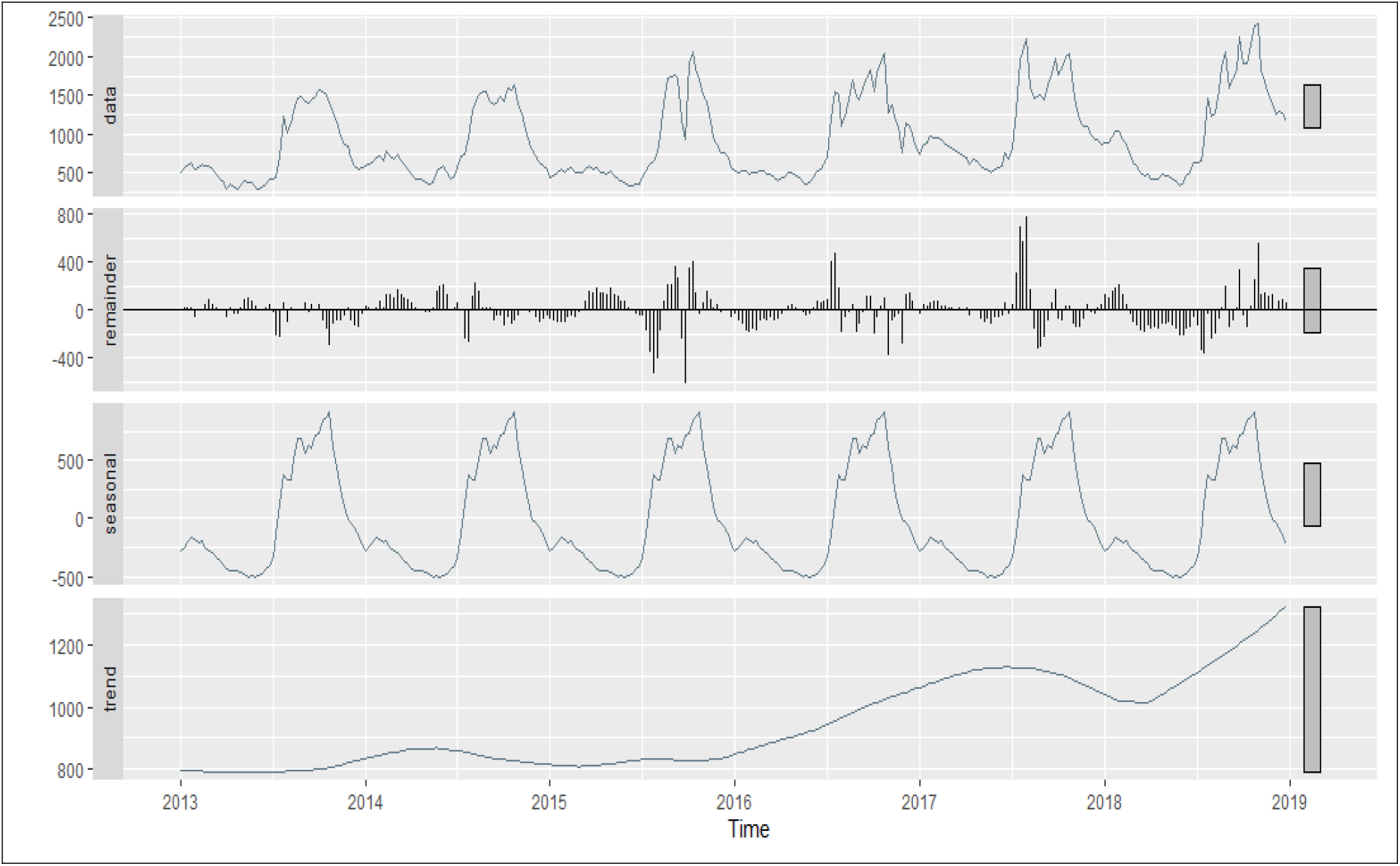
Time series of incidence and its seasonal and trend components. Raw time series (data panel). Time series noise (remainder panel). Time series seasons (seasonal panel). Time series trend (trend panel)

A more detailed analysis of the time series using the Change point method allowed us to characterize distinct seasonal dynamics in these two phases.

The stable phase 1 (2013-2016) exhibited only two alternating transmission periods: low from January to July (included) and high from August to December (included).

During increasing phase 2, in 2017 to 2018, an intermediate transmission period from mid-November to early January intercalated between the low transmission period from mid-February to early June and the high transmission period from July to late December (Figure 6).

**Figure 6:**
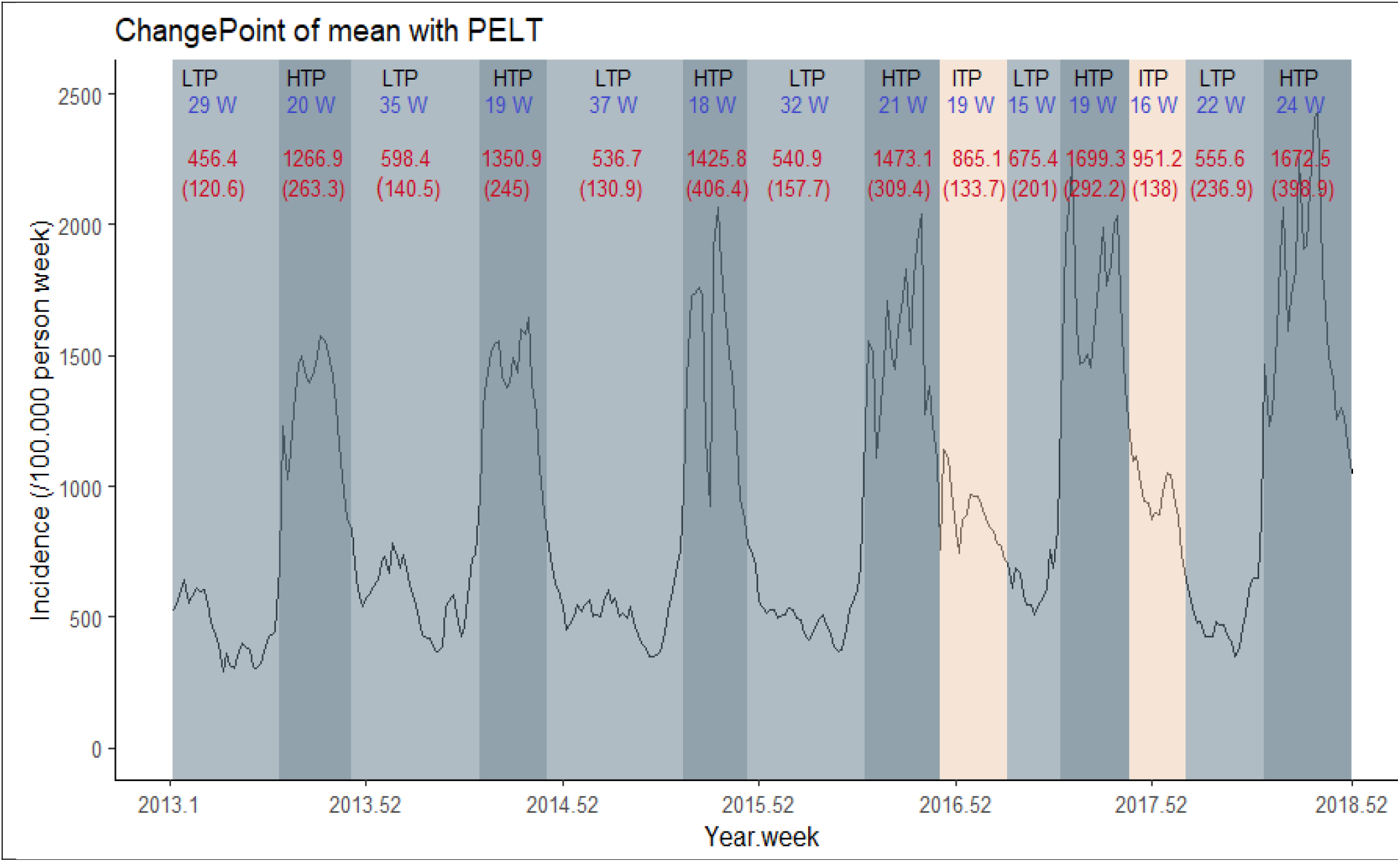
Trends in weekly malaria incidence and transmission periods from 2013 to 2018 with their duration (weeks, in blue) and the averages of the incidence rates with their standard deviation (red numbers). LTP: Low transmission period; HTP: High transmission period; ITP: Intermediate transmission period.

We have previously (Figure 2) observed that the incidence of malaria had an increasing trend over the years. The mortality rate has a decreasing trend over the same period (Figure 7).

**Figure 7:**
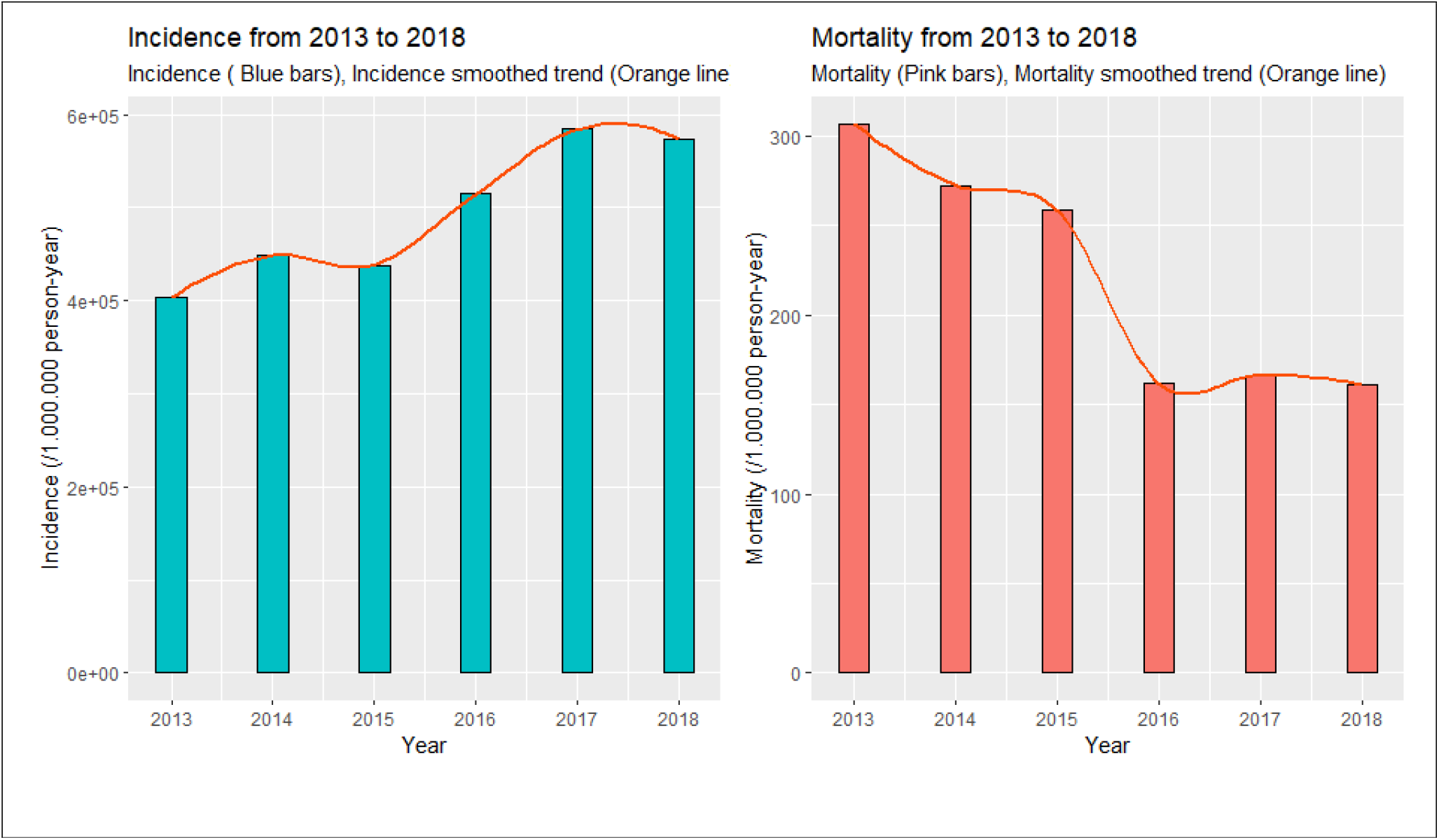
Incidence and Mortality by Year

### Spatial mapping

Choropleth maps of malaria incidence showed the evolution and distribution of malaria incidence across the country.

The health districts most affected over the study period vary from one year to another; from 2013 to 2015, the most affected districts were located in the center, central-eastern and some districts in the north with at least 45,000 cases per 100,000 persons/year. By 2016, all districts except those located in the central-western and the Sahel regions were affected with at least 45,000 cases per 100,000 persons/year, but with a particularly pronounced incidence in the southwest and in the east with at least 62,000 cases per 100,000 persons/year. From 2017 to 2018 all districts except those in the Sahel region were affected with at least 45,000 cases per 100,000 persons/year but with a particularly pronounced incidence in the southwest, south, central-east and east with at least 62,000 cases per 100,000 persons/year. For this period (2017-2018), some districts in the south-west and the east reached 95000 cases per 100,000 persons/year. In all cases, the Sahel and the central-western region are the least affected regions. In addition, from 2016 onwards, the incidence maps “light up” showing an increase in malaria cases in Burkina Faso (Figure 8).

**Figure 8:**
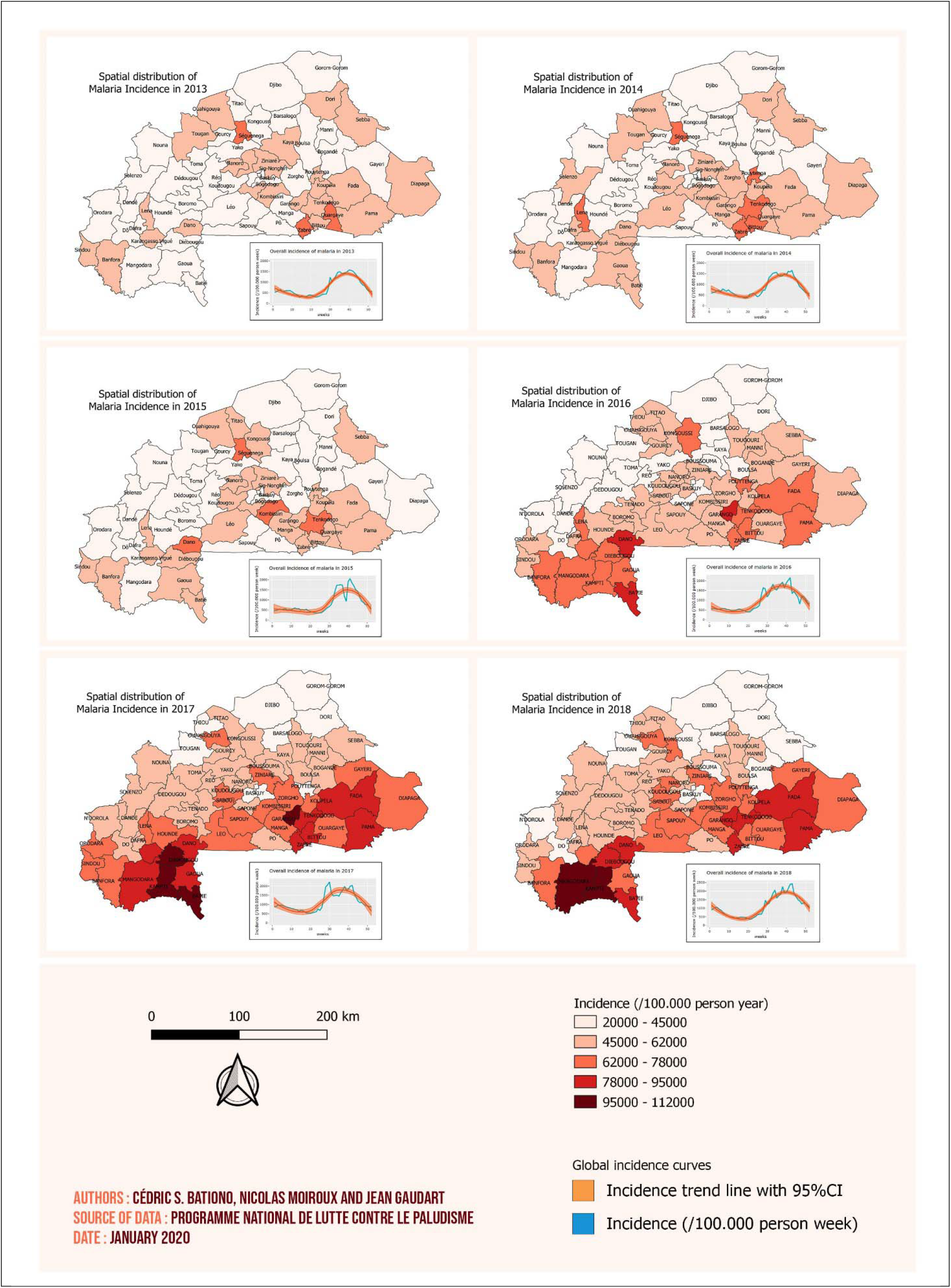
District level malaria incidence maps from 2013 to 2018. Each map has on the bottom right the overall smoothed malaria incidence time series with the 95% confidence interval.

Maps of mean rainfall, temperature and normalized difference vegetation index data over the entire study period are shown in figures 9, 10 and 11. For the rainfall and the NDVI data, we observed a north-south gradient with the highest rainfall and NDVI in south-west, south, central-eastern and southeastern regions. The mean temperature has the same trend with the smallest temperatures localized in the south-west.

**Figure 9:**
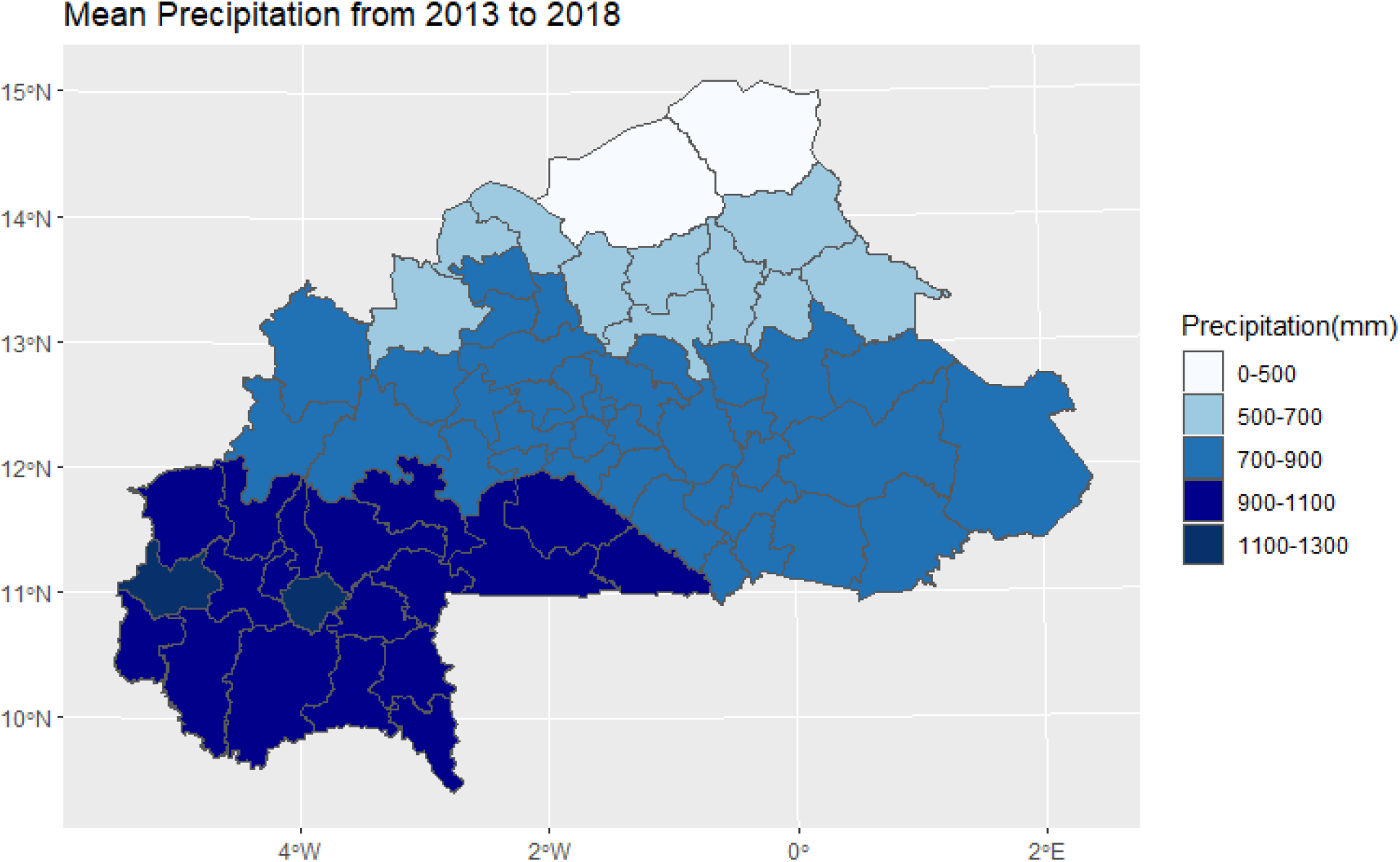
Mean rainfall map from 2013 to 2018.

**Figure 10:**
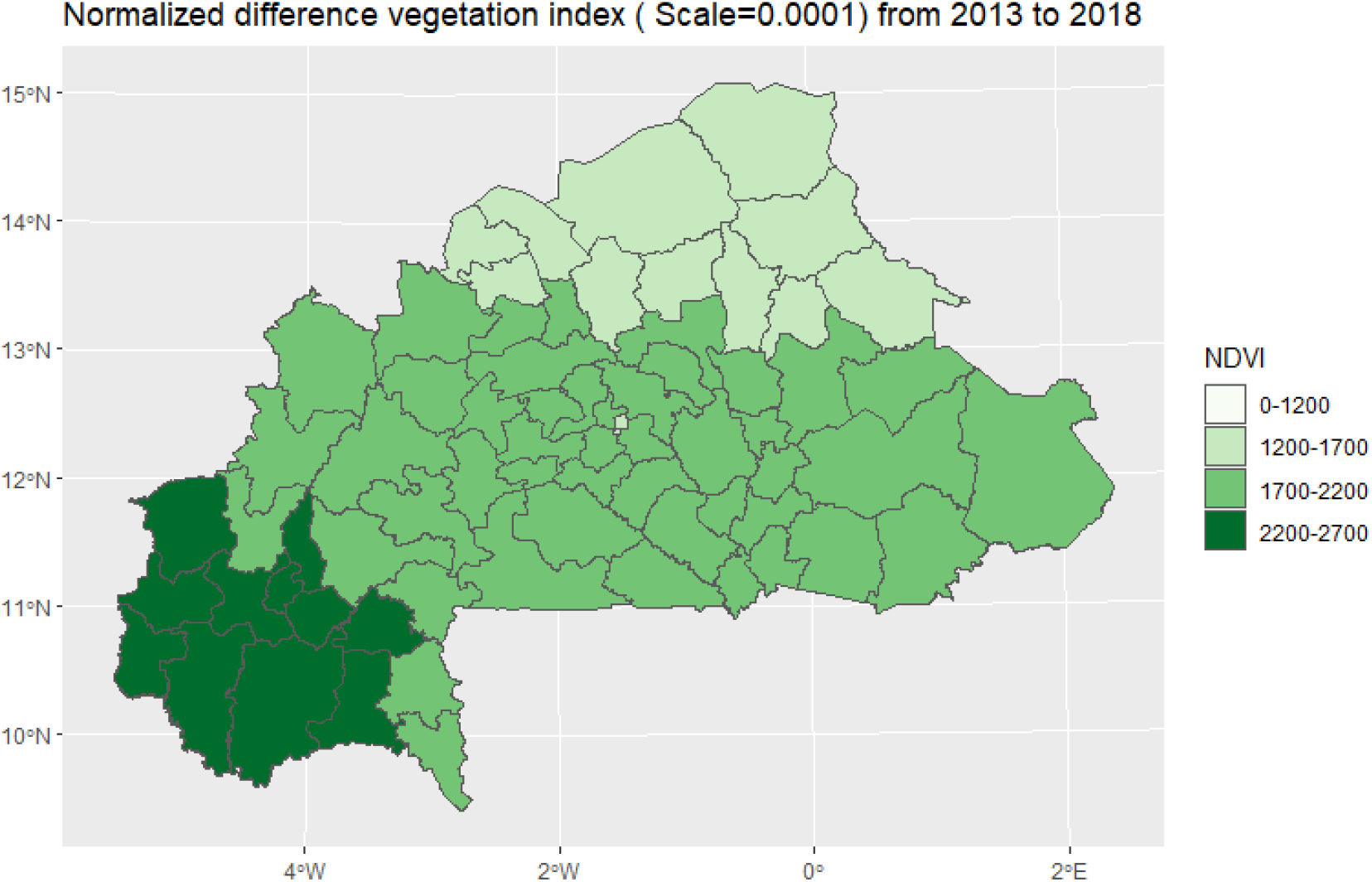
Mean NDVI map from 2013 to 2018.

**Figure 11:**
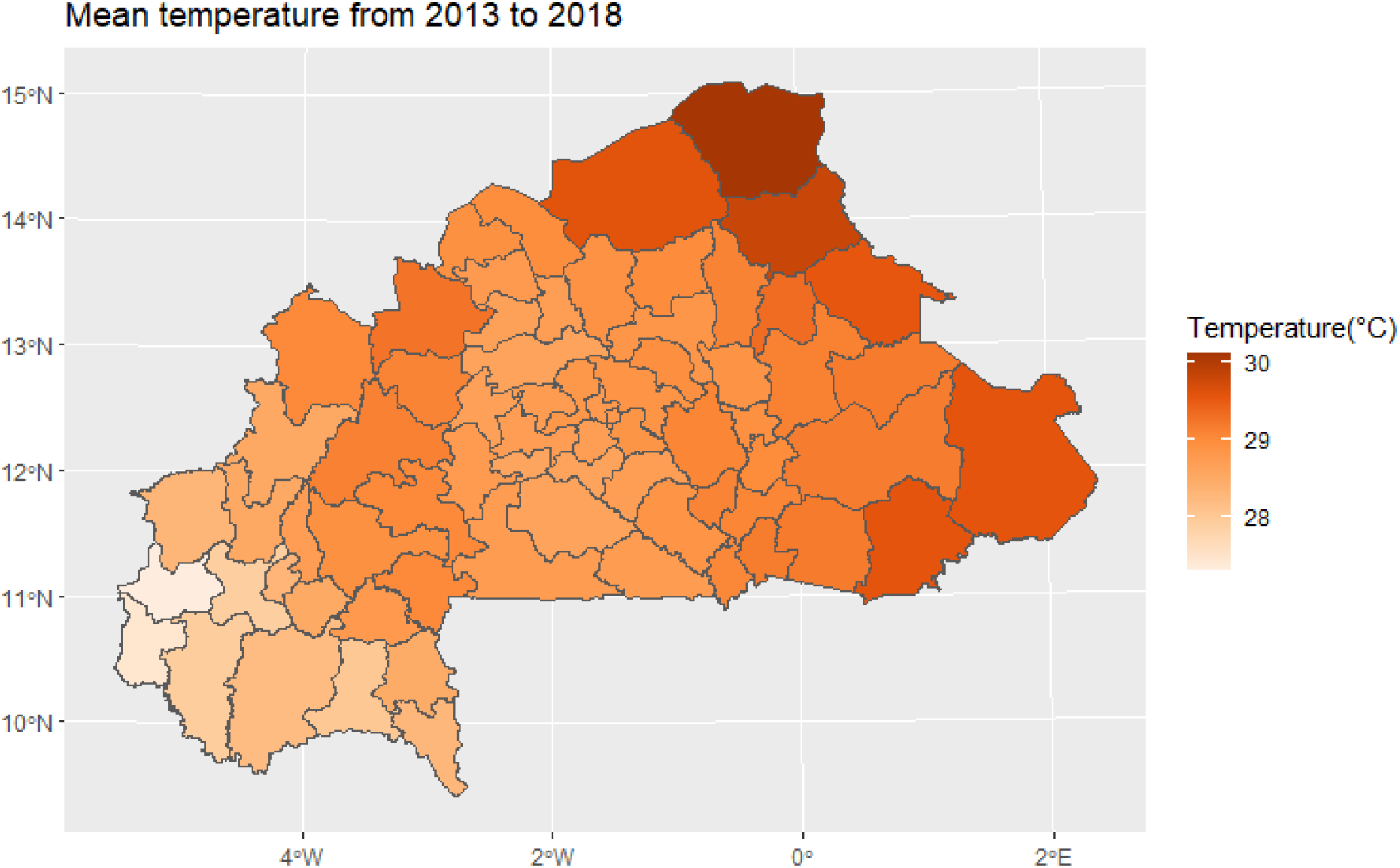
Mean temperature map from 2013 to 2018.

## Discussion

We observed a nearly stable incidence over the period 2013 to 2015 through years and across the country. An increase in malaria incidence from 2016 to 2018 was observed associated to an extension of the most affected areas. Malaria incidence in Burkina Faso had a constant seasonality superimposed on periods of high rainfall with a lag even if throughout each year there are periods of low transmission and even intermediate transmission from 2017 onwards (Figure 6).

This increase in the incidence may be due to a better access to diagnosis and treatment with rapid diagnosis tests (RDTs) and/or to a better surveillance and information system (thus capturing a higher proportion of malaria cases treated in health centers).

Indeed, an efficient monitoring and information system gives the advantage of continuously collecting almost exhaustive data in each district. For most districts, these data are the only readily available source of malaria information that program managers can use ^31,32^.

Efficient surveillance systems show regular seasonal variations in case numbers, coinciding with transmission patterns. They also show decreases in morbidity and mortality following interventions and can alert administrators to unexpected increases ^31,33^. During the study period, Burkina Faso made efforts to improve access to care and/or the surveillance/data collection system, including the introduction of a second long lasting insecticidal nets (LLINs) distribution campaign in 2013, the coverage of seasonal chemoprevention campaigns, which increased from 7 to 65 health districts between 2013 and 2018, and the introduction of intermittent preventive treatment during pregnancy at the community level.

On the other hand, an increase in the incidence could also mean that malaria is escaping prevention and control policies and strategies. This could be caused by inadequate implementation of malaria control and prevention policies, particularly the distribution of seasonal malaria chemoprevention (SMC) and intermittent preventive treatment (IPT) to pregnant women at the wrong time, difficulties (stocks out) in accessing rapid diagnostic tests (RDTs) and artemisinin-based combination therapies (ACTs), and the lack of information on the use and efficient storage of LLINs may hinder the reduction of malaria cases because they do not have a protective effect on the population ^34–39^.

It may also be that vectors have been resistant to insecticides or parasites to treatment; indeed, in the past decade, pyrethroids resistance in major malaria vectors in Sub-Saharan Africa (*Anopheles gambiae* (including *An. gambiae sensu stricto* (s.s.) and *An. coluzzii*), *An. arabiensis* and *An. funestus* s.s.) has spread across the continent being prevalent in west ^40^. The pyrethroids resistance is a great concern because pyrethroids are the main insecticide class recommended for long lasting insecticidal nets impregnation ^41^. Resistance to long lasting insecticidal nets exposure increases mosquito survival, which may lead to rising malaria incidence and fatality ^42^. Nevertheless, parasite resistance to treatment seems unlikely, when most studies conducted between 2010 and 2017 show that ACTs remain effective, with overall efficacy rates above 95% in the WHO African regions ^43^.

In addition to that, the onset of the intermediate transmission period may be contributing to an increase in cases due to a slower decline in incidence.

Another unexplored avenue that could explain this almost linear increase is that, from December 2013 to march 2016 it was the outbreak of hemorrhagic fever Ebola virus which raged in West Africa and which was the largest and most complex since the discovery of the virus in 1976 ^44^. During this period, Burkina Faso mobilized efforts for prevention and response against Ebola ^45^, reducing efforts against other diseases such as malaria.

Malaria incidence in Burkina Faso had a constant seasonality superimposed on periods of high rainfall. Contrary to this finding, low temperatures were superimposed on the highest incidences. Indeed, above a certain temperature (>34°C), the development of Anopheles larvae is inhibited and thus reduces the survival of adult Anopheles ^46^. Low temperatures therefore favor the survival of larvae (above a minimal temperature threshold of 18°C)

The decrease in malaria mortality from 2013 to 2018, during which period the incidence gradually increased (Figure 7), can be explained by a better management of malaria cases. Indeed, following the increase in resistance to conventional treatment (chloroquine) ^47–49^), the WHO has in 2006 made recommended to rely on artemisinin-based drugs for the management of both uncomplicated and severe falciparum malaria cases^50^. In Burkina Faso, this strategy is supported by the recommendations of the NMCP, which recommends the following management strategies ^51^: early case management in health facilities and at the community level, with particular emphasis on children aged 3 to 59 months since 2009 ^52^, intermittent preventive treatment (IPT) for pregnant women since 2017, universal access to rapid diagnostic tests (RDTs) since 2009 and artemisinin-based combination therapies (ACTs) since 2006, and seasonal chemio-prevention (SCP) for children under 5 since 2013.

In any case, the health system does not capture all cases and deaths (only cases and deaths in health facilities are captured).

It is therefore imperative to conduct studies on these cases not captured by the system and especially to develop national approaches to take them into account while analyzing data for decision making.

Spatial analysis has shown a heterogeneous distribution of malaria incidence. The most affected districts (located in the south, southeast, and east) showed little change from 2013 to 2018, although the intensity of infection in these areas increased during this period. This heterogeneity can be explained by the fact that the health districts are spread across different climatic zones. As weather is a factor known to influence malaria incidence ^7,26,27^, different climatic zones may be affected differently by malaria. In addition, a number of studies have found an association between spatial inequalities in access to health care and spatial heterogeneity in malaria incidence ^53,54^.

Also, entomological factors (heterogeneous vector abundance) and other potential explanatory factors should be investigated, including socioeconomic factors (level of education, income, professional activity, individual and societal behavior, etc…)^55^and factors related to LLIN use ^56^.

## Conclusion

In this epidemiological study, we assessed malaria incidence and mortality in Burkina Faso. We compared the different trends of malaria incidence over the period 2013-2018. This work highlights malaria incidence increase and extension through years and across the country, together with a mortality reduction, suggesting an important role of the improvement of access to diagnostic and treatment. Malaria surveillance appears now challenged by other epidemics of acute fever diseases such as dengue, which have to be identified and separated from malaria. Regular analysis of spatio-temporal data is one of the keys to improving the understanding of malaria dynamics leading to better preparedness and reactivity of malaria control policies.

## Data Availability

The datasets analyzed in this study may be available from the last author on reasonable request.

## Acknowledgements

The authors would like to thank the Ministry of Health of Burkina Faso, in particular the National Malaria Control Program and the Health Information Systems Department for facilitating data collection.

## Contributions

C.S.B and J.G designed the study; C.S.B., J.G., and N.M. designed the statistical analysis plan; C.S.B. conducted the statistical analysis under the supervision of J.G. and J.L.; C.S.B. performed the cartographic analysis with the participation of M.C; C.S.B, V.L, B.S, J.L and J.G validated and interpreted the results; C.S.B., J.G., and N.M. wrote the manuscript; all authors read and approved the final manuscript.

